# Stabilization of the coronavirus pandemic in Italy and global prospects

**DOI:** 10.1101/2020.03.28.20045898

**Authors:** Igor Nesteruk

**Affiliations:** Institute of Hydromechanics, National Academy of Sciences of Ukraine, Zheliabova St, 8/4, UA-03680 Kyiv, Ukraine; National Technical University of Ukraine “Igor Sikorsky Kyiv Polytechnic Institute”, Peremohy Av, 37, UA-03056, Kyiv, Ukraine

**Keywords:** coronavirus pandemic, epidemic outbreak in Italy, coronavirus COVID-19, coronavirus 2019-nCoV, mathematical modeling of infection diseases, SIR model, parameter identification, statistical methods

## Abstract

The pandemic caused by coronavirus COVID-19 are of great concern. A detailed scientific analysis of this phenomenon is still to come, but now it is urgently needed to evaluate the parameters of the disease dynamics in order to make some preliminary estimations of the number of cases and possible duration of the pandemic. The corresponding mathematical models must be simple enough, since their parameters are unknown and have to be estimated using limited statistical data sets. The SIR model, statistical approach to the parameter identification and the official WHO daily data about the confirmed cumulative number of cases were used to calculate the SIR curves and make some estimations and predictions. New cases in Italy could stop to appear after May 12, 2020, and the final number of such accumulated cases could be around 112 thousand. Some prospects for the global pandemic dynamics are discussed.

## Introduction

Here, we consider the development of epidemic outbreak in Italy caused by coronavirus COVID-19 (2019-nCoV) (see e.g., [1]). Some investigations of the epidemic spreading in mainland China [2–7] could be useful to understand the epidemic outbreak in other countries, since we deal with the same pathogen. A preliminary comparison of the epidemic dynamics in Italy and in mainland China has been done in [8, 9]. In [10] the global coronavirus epidemic dynamics was analyzed. In this paper we will use the official WHO daily data [1] for the confirmed accumulated number of patients (victims) *V(t)* (number of persons who caught the infection and got sick; *t* is time measured in days), the SIR model [11–14] and the statistics-based method of parameter identification [14] in order to calculate the pandemic characteristics and to make some estimations and predictions.

### Data

The official data about the accumulated number of confirmed cases in Italy *V*_*j*_; European region *V*_*Ej*_, USA *V*_*Uj*_ and global numbers *V*_*Gj*_ (without cases inmainland China and the Republic of Korea) from the WHO daily situation reports (numbers 33-65, [1]) will be used. The corresponding moments of time *t*_*j*_ (in days) are also shown in Table 1.

**Table 1.** Official cumulative numbers of confirmed cases in Italy, European region, USA and World (without mainland China and South Korea), [1].

| Day in February<br>and March, 2020 | Time moments<br>in days $t_j$ | Number of cases<br>in Italy, $V_I$ | European<br>Region, $V_{EJ}$ | USA, $V_{Uj}$ | Global without<br>cases in mainland<br>China and South<br>Korea, $V_{GJ}$ |
| --- | --- | --- | --- | --- | --- |
| 21 | -1 | 9 | 54 | 35 | 1056 |
| 22 | 0 | 76 | 121 | 35 | 1167 |
| 23 | 1 | 124 | 169 | 35 | 1306 |
| 24 | 2 | 229 | 279 | 53 | 1482 |
| 25 | 3 | 322 | 379 | 53 | 1657 |
| 26 | 4 | 400 | 486 | 59 | 1898 |
| 27 | 5 | 650 | 798 | 59 | 2354 |
| 28 | 6 | 888 | 1107 | 62 | 2859 |
| 29 | 7 | 1128 | 1457 | 62 | 3433 |
| 1 | 8 | 1689 | 2136 | 62 | 4562 |
| 2 | 9 | 2036 | 2732 | 64 | 5753 |
| 3 | 10 | 2502 | 3367 | 108 | 7341 |
| 4 | 11 | 3089 | 4307 | 129 | 8993 |
| 5 | 12 | 3858 | 5820 | 148 | 11197 |
| 6 | 13 | 4636 | 7491 | 213 | 14347 |
| 7 | 14 | 5883 | 9453 | 213 | 17593 |
| 8 | 15 | 7375 | 12242 | 213 | 21291 |
| 9 | 16 | 9172 | 15130 | 472 | 25265 |
| 10 | 17 | 10149 | 18124 | 696 | 29609 |
| 11 | 18 | 12462 | 23112 | 987 | 36410 |
| 12 | 19 | 15113 | 28892 | 1264 | 43788 |
| 13 | 20 | 17660 | 36263 | 1678 | 53427 |
| 14 | 21 | 21157 | 45060 | 1678 | 64304 |
| 15 | 22 | 24747 | 55623 | 1678 | 78197 |
| 16 | 23 | 27980 | 64135 | 3503 | 89680 |
| 17 | 24 | 31506 | 74703 | 3536 | 101612 |
| 18 | 25 | 35713 | 86994 | 7087 | 120173 |
| 19 | 26 | 41035 | 104425 | 10442 | 144121 |
| 20 | 27 | 47021 | 128434 | 15219 | 175858 |
| 21 | 28 | 53578 | 151173 | 15219 | 201747 |
| 22 | 29 | 59138 | 171226 | 31573 | 242368 |
| 23 | 30 | 63927 | 195261 | 42164 | 281973 |
| 24 | 31 | 69176 | 220249 | 51914 | 323194 |

### SIR model and optimal values of its parameters

The SIR model for an infectious disease [6, 11–14] relates the number of susceptible persons *S* (persons who are sensitive to the pathogen and **not protected**); the number of infected is *I* (persons who are sick and **spread the infection**; please don’t confuse with the number of still ill persons, so known active cases) and the number of removed *R* (persons who **no longer spread the infection**; this number is the sum of isolated, recovered, dead, and infected people who left the region); *α* and *ρ* are constants.

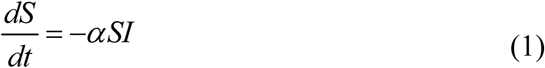

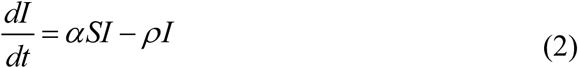

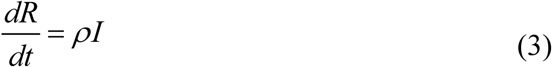

To determine the initial conditions for the set of equations (1–3), let us suppose that at the moment of the epidemic outbreak *t*_0_, [6, 14]:

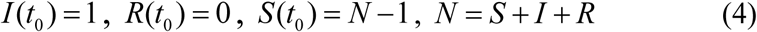

The analytical solution for the set of equations (1–3) was obtained by introducing the function *V*(*t*) = *I*(*t*) + *R*(*t*), corresponding to the number of victims or cumulative confirmed number of cases, [14]:

The solution for the SIR set of differential equations depends on four parameters *N, α, ν* = *ρ* / *α, t*_0_, which can be identified with the use of the statistical approach developed in [14]. This method and *V*_*j*_ data set for Italy were used to define the optimal (the most reliable) values of four parameters and calculate numbers of infected *I*, susceptible *S*, removed *R* persons and the number of victims *V=I*+*R*. Corresponding dependences versus time are shown in Fig. 1.

**Fig. 1.**
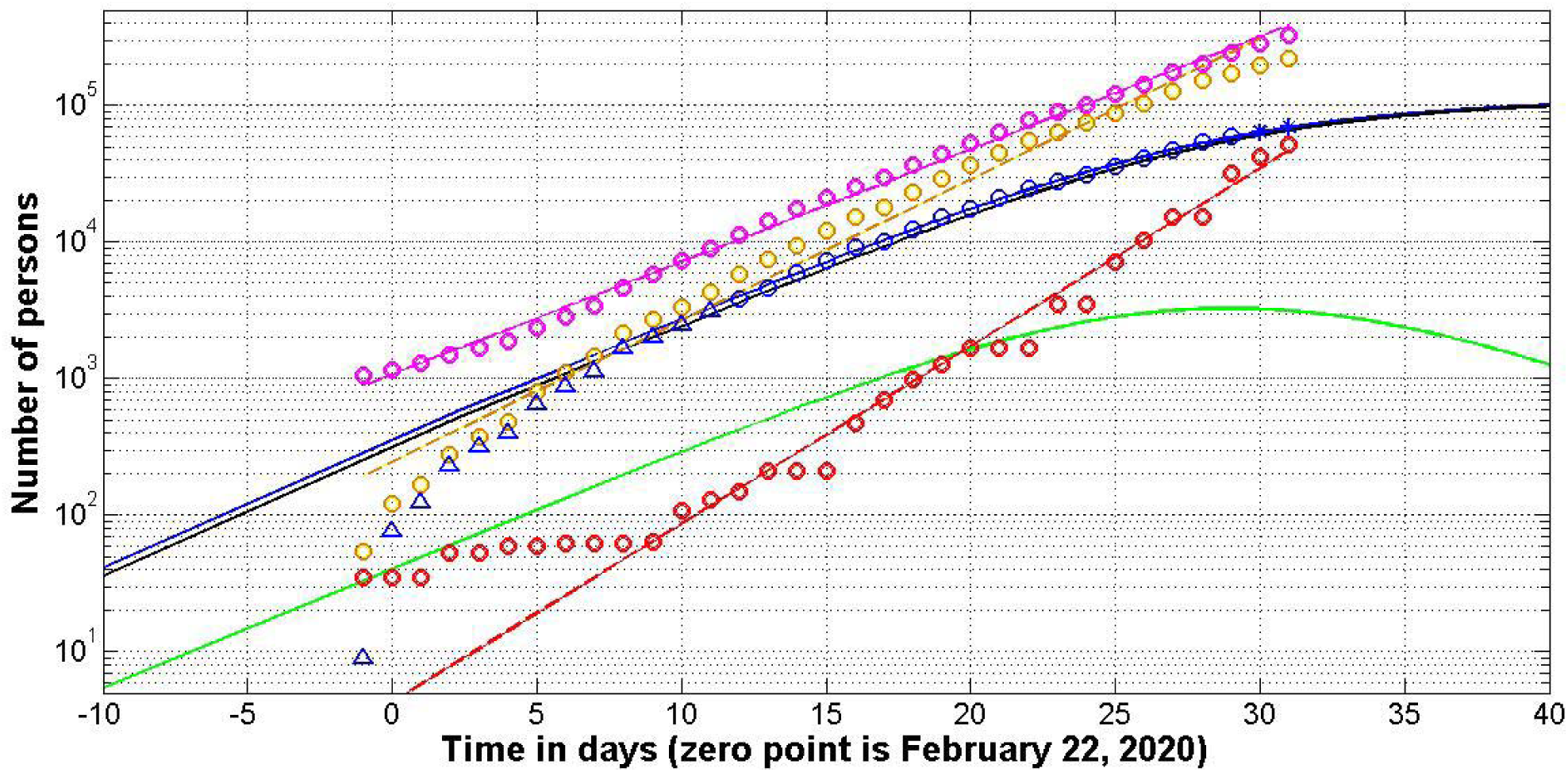
SIR curves for Italy (solid lines). Pandemic development in Europe, USA and in the World (without cases in mainland China and South Korea) (dashed lines). For Italy: numbers of infected *I* (green line), removed *R* (black line) and the number of victims *V=I*+*R* (blue line); “circles” correspond to the confirmed accumulated number of cases taken for calculations; “triangles” correspond to the cases during initial stage of the epidemic; “stars” –last two data points used only for a verification of the prediction. Brawn, red and magenta markers represent respectively the numbers of cases in European region, USA and in the World (without cases in mainland China and South Korea); corresponding dashed lines fit the points.

### Results for Italy

Usually the number of cases during the initial period of an epidemic outbreak is not reliable. To avoid their influence on the results, only *V*_*j*_ values for the period February 5-22, 2020 (12 ≤ *t*_*j*_ ≤ 29) were used for calculations (see blue “circles” in Fig. 1). Other points were used only for comparison (blue “triangles”) and verification of predictions (blue “stars”). The use of data corresponding the initial stage of the epidemic (“circles” and “triangles” together) did not yield a stable reliable prediction. The calculated optimal values of parameters are:

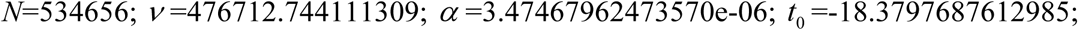

Now every person familiar with differential equations can use this data to integrate Eqs. (1–3) with initial conditions (4) to obtain the SIR curves and to check the results of calculations (it is also possible to use the analytical solution available in [6]). The values of final number of susceptible persons *S*_∞_ ≈ 423108 and the final number of victims (final accumulated number of cases) *V*_∞_ ≈ 111548 were calculated. Unfortunately, in Italy more people will be infected in comparison with mainland China, where the saturation level *V*_∞_ ≈ 81257 was predicted in [9] on March 4, 2020 in [9] (on March 24 the accumulated number of cases confirmed in mainland China is 81848, see [1], situation report No 65). On March 9, 2020 it was calculated that the epidemic in Italy develops more rapid than it was in China, [15]. Unfortunately, this conclusion seems to be true.

To estimate the duration of the coronavirus epidemic outbreak in Italy, we can use the condition *V* (*t* _*final*_) *=*1 which means that after this moment less than one person still spread the infection. The calculations give us the value *t_final_* ≈ 79.5. According to this estimation, we can expect that local transmission of the epidemic in Italy could stop only after May 12, 2020, provided that existing quarantine measures and patient isolation rates continue.

It is also possible to calculate the value of parameter *ρ* = *να* =1.6564 and the inverse value 1/ *ρ* **=**0.6037. Thus, the average time of spreading the infection in Italy can be estimated as 14.5 hours. By comparison, in South Korea was approximately 4.3 hours [16] and in mainland China - 2.5 days [9]. By mid-April 2020, there will still be more than thousand people spreading the infection in Italy (see green line in Fig. 1).

The calculated value *t*_0_ and blue line in Fig. 1 demonstrate that the first cases of coronavirus infection have not been identified in Italy and sick people spread it rather long time. As a result, more cases are expected in Italy in comparison with China. Probably, due to the rapid isolation of infected persons, the more or less stable number of cases in South Korea (9137 on March 24) is much smaller than in Italy.

All the parameters in SIR model are supposed to be constant. If the quarantine measures and speed of isolation change or new infected persons are coming in the country, the accuracy of the prediction reduces.

### Discussion of global prospects

Since the recent situation in mainland China and the Republic of Korea is stable, The estimations of global pandemic prospects have been done without cases in this two regions. The corresponding numbers *V*_*Gj*_ are shown in the last column of Table 1 and in the Fig. 1 (magenta “circles”). The number of cases *V*_*Ej*_ and *V*_*Uj*_ are also shown in Fig. 1 (brown and red “circles” respectively). It can be seen that *V*_*Ej*_, *V*_*Uj*_ and *V*_*Gj*_ numbers follow straight lines in the logarithmic scale. It means that the epidemic dynamics in these regions is still exponential and is far from stabilization.

To estimate the slopes of these lines the linear regression for the values log(*V*_*Yj*_), *Y* = *E,U, G* was used (see, e.g., [14, 17]). The corresponding best fitting dashed lines are shown in Fig.1. To avoid influence of the initial epidemic outbreak stages, calculations have been done only for *V*_*Yj*_ > 100, *Y* = *E,U*; repeating values were used only once. It can be seen that the most rapid is the epidemic in USA (the number of cases duplicates every 2.31 days). The European and global duplications rates are 2.91 and 3.65 days respectively. Without stabilization by mid-April 2020 (22 days after March 24) we can have in Europe 41 million cases; 38 millions in USA and 211 millions globally. Let us hope that quarantine measures and fast isolation of infected persons will reduce these sad figures.

## Data Availability

data sets are in the text

## Acknowledgements

I would like to express my sincere thanks to Gerhard Demelmair and Ihor Kudybyn for their help in collecting and processing data.

